# Improving Efficiency in the Inpatient Echocardiography Laboratory: A Quality Improvement Project

**DOI:** 10.1101/2025.01.28.25321298

**Authors:** Stefani Samples, Zhanna Roytman, Carrie McCaw, Pei-Ni Jone

## Abstract

**Background:** Pediatric echocardiograms are performed daily on inpatient hospital units, predominantly in the Cardiac Intensive Care Unit. Understanding the precise study indication from the ordering providers is critical for completing imaging. Given the volume of echocardiograms ordered daily and staffing shortages, timely completion of echocardiograms is important for maintaining efficiency and productivity. The aim of this study was to identify areas to improve efficiency using quality improvement methodologies.

**Methods:** The SMART aim was to decrease the total echo completion time by 10% within 6 months. Using a fishbone diagram, limitations in the ordering system and inadequate ordering provider knowledge were identified as areas for improvement. Primary process and outcome metrics included total echo completion time, pre-echocardiogram buffer time, and order documentation of pertinent clinical history. PDSA 1 implemented order updates to provide increased specificity in orders with a picklist of common reasons for inpatient echocardiograms. PDSA 2 implemented a physician alert for STAT studies to reduce the number of inappropriate orders. PDSA 3 implemented additional order updates and staff education.

**Results:** Total echo completion time decreased with each cycle for total 25% reduction. This correlated with a decrease in the pre-echocardiogram buffer time of 56% without any significant changes in the number of images obtained per study or the daily inpatient volume. The documentation of pertinent clinical history increased by 18% and maintained at 99% appropriate documentation for the final two PDSA cycles. Pre and post intervention surveys indicated an overall improvement in the ordering process for both the inpatient ordering providers and echocardiography laboratory staff.

**Conclusions:** Echocardiography laboratory efficiency significantly improved throughout the course of this project without negative effects on the ordering providers or echocardiography laboratory staff. Our project demonstrates the ability to affect clinically relevant improvements utilizing simple workflow process changes in the pediatric echocardiography laboratory.

## Introduction

Echocardiography laboratories (echo lab) provide imaging services which play a crucial role in the diagnosis and management of congenital heart disease. Pediatric echocardiograms can be performed for a complex set of reasons to evaluate one or multiple aspects of cardiac anatomy or physiology. They are performed on both outpatients and inpatients at pediatric hospitals across the country. Each echocardiogram can routinely take 45-60 minutes, and imaging time can become extended with patient related issues or advanced imaging requirements such as 3-dimensional echocardiography. Creating an efficient workflow is critical to ensure appropriate throughput of patients throughout the workday. Despite this, there are limited previous studies centering on the efficiency of the pediatric echo lab, with many studies only addressing efficiency peripherally or from the patient’s point of view. ^1-3^

There are multiple different hospital departments interacting through the echocardiogram ordering process, including the various intensive care unit (ICU) staffs, resident physicians, and echo lab staff. Given this, there can be many interrelated contributors to inefficiency (Figure 1). At our institution, we noticed a pattern of inefficiencies in our inpatient echo lab creating a slowing in our daily workflow and making it difficult to complete all the inpatient echocardiograms ordered on many days. With these inefficiencies, echocardiograms could routinely be delayed until the following day or would be completed in the evenings by the on-call fellows who are less experienced at performing complex echocardiograms.

**Figure 1:**
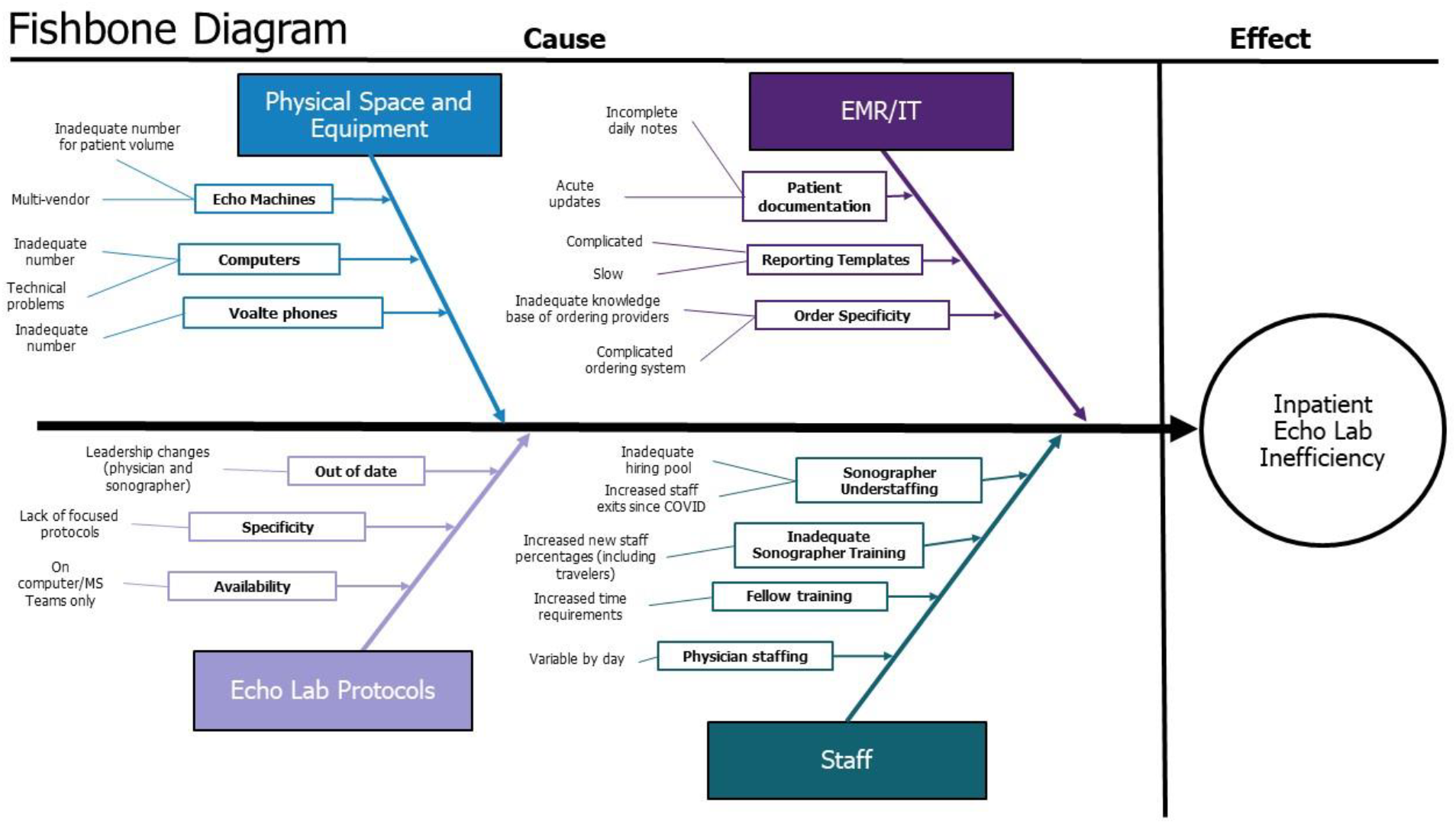
Fishbone diagram detailing potential causes of inpatient echocardiography laboratory inefficiency.

As a result of these observations, a quality improvement (QI) project was started with an aim of improving the inpatient echo lab efficiency by decreasing the time needed for each echocardiogram to be completed by 10% within 6 months.

## Methods

### Context

At our institution, we have 22 pediatric cardiac sonographers performing echocardiograms throughout the day between our main hospital campus and multiple satellite clinic locations. The echo lab performs over 19,000 echocardiograms per year, which routinely means 80 to 100 echocardiograms are performed between the outpatient and inpatient imaging areas each weekday. As a result, we have separate inpatient and outpatient echocardiogram reading workspaces to prioritize the relevant imaging in each area. There is a dedicated inpatient echo physician and another 2-4 outpatient echo physicians staffing these labs respectively daily. Inpatient echocardiograms can be ordered and completed 24/7, however the primary concentration is during the day on a normal weekday. In the fourth quarter of 2022, our echo lab performed an average of 16.5 inpatient studies each weekday. The majority of our inpatient studies are performed in the various ICU settings at our hospital, Cardiac (CCU), Neonatal, and Pediatric ICUs, with an average of 7.9 studies performed in our CCU each weekday.

The process of completing an inpatient echocardiogram has multiple steps for the echo lab staff (Figure 2). Each sonographer reviews the echo order before going to the patient room to complete imaging and completes any preparatory work needed. This study prep time is also the initial buffer time which is variable. After initial imaging is completed, each echocardiogram is reviewed by an attending physician prior to the sonographer leaving the room to ensure no additional imaging is required. Once imaging is completed, the sonographer prepares a preliminary report in our reporting system, Syngo Dynamics (Siemens Healthcare, USA), and the attending physician reviews, edits, and finalizes each report.

**Figure 2:**
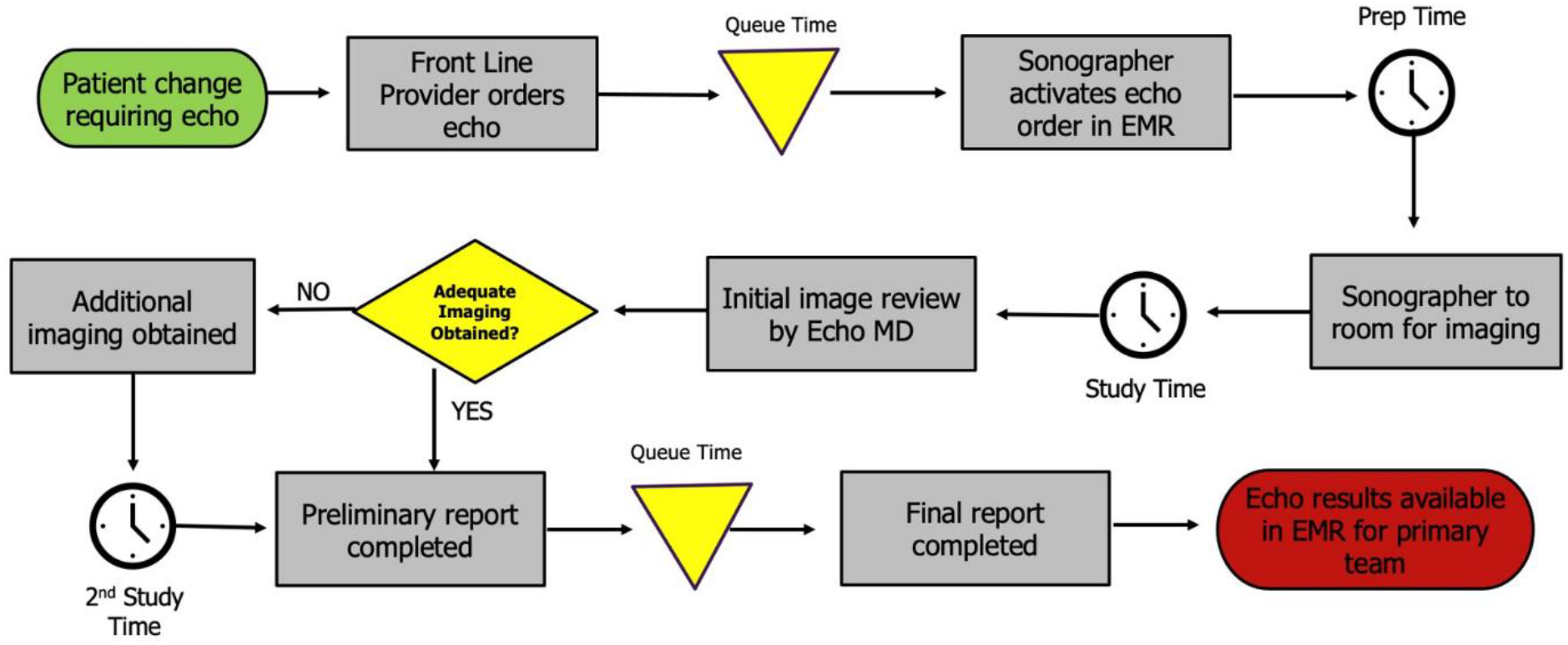
Process map demonstrating steps from ordering an echocardiogram through completion of imaging until finalization of report.

The existing ordering system was through the electronic medical record system Epic (Epic Systems Corporation, USA) with a single comments area designated for ordering providers to provide the clinical history or reason for the echo to guide the echo lab staff on the area of clinical concern. These order comments are automatically transmitted to the “Indication” field in our echocardiogram reports in Syngo Dynamics (Siemens Healthcare, USA). These order comments were generally vague and often similar across multiple patients’ echo orders which required the performing sonographer or attending physician to extensively review the patients’ medical records for clarity and/or speak with the primary team for additional clinical details in order to ensure all appropriate imaging was obtained. All of these steps added significant time to the workflow for each echocardiogram each day, thereby making it more difficult to perform the same number of echocardiograms each day. STAT echocardiogram orders being placed inappropriately also slowed the daily workflow. While the primary medical teams generally desired these echocardiograms to be performed sooner, they were not usually emergent. The placement of these STAT orders required the echo lab team to stop other work and investigate if these echocardiograms were truly emergent, thereby slowing the daily workflow further.

### Intervention

Stakeholder conversations with the CCU and echo lab staff were held to determine the common pain points when ordering or completing echocardiograms. A baseline staff survey was utilized for more extensive feedback with questions guided by those initial stakeholder conversations. From this combined feedback, we developed a key driver diagram incorporating our SMART (specific, measurable, achievable, relevant, and time-bound) aim (Figure 3). Given the relatively large volume and turnover of staff in all areas, modification of the echo order to promote additional specificity was determined to be the primary intervention. Echocardiogram orders over a 3 months period were reviewed for the reason for study, and a comprehensive list was developed. Working with our information technology (IT) team, picklists were developed and inserted into the echocardiogram orders. Two picklists were developed total, one for inpatient orders and another for outpatient orders, given the differences in common reasons for study in these areas. The selections in the picklists were put in order of frequency of use, with the expected most used reasons for study located at the top of each list (Figure 4). Additional refinement and additions to the list were incorporated during the project. Testing was performed with each update to validate continued transmission of this field to our echocardiogram reports in Syngo Dynamics (Siemens Healthcare, USA) to ensure appropriate documentation.

**Figure 3:**
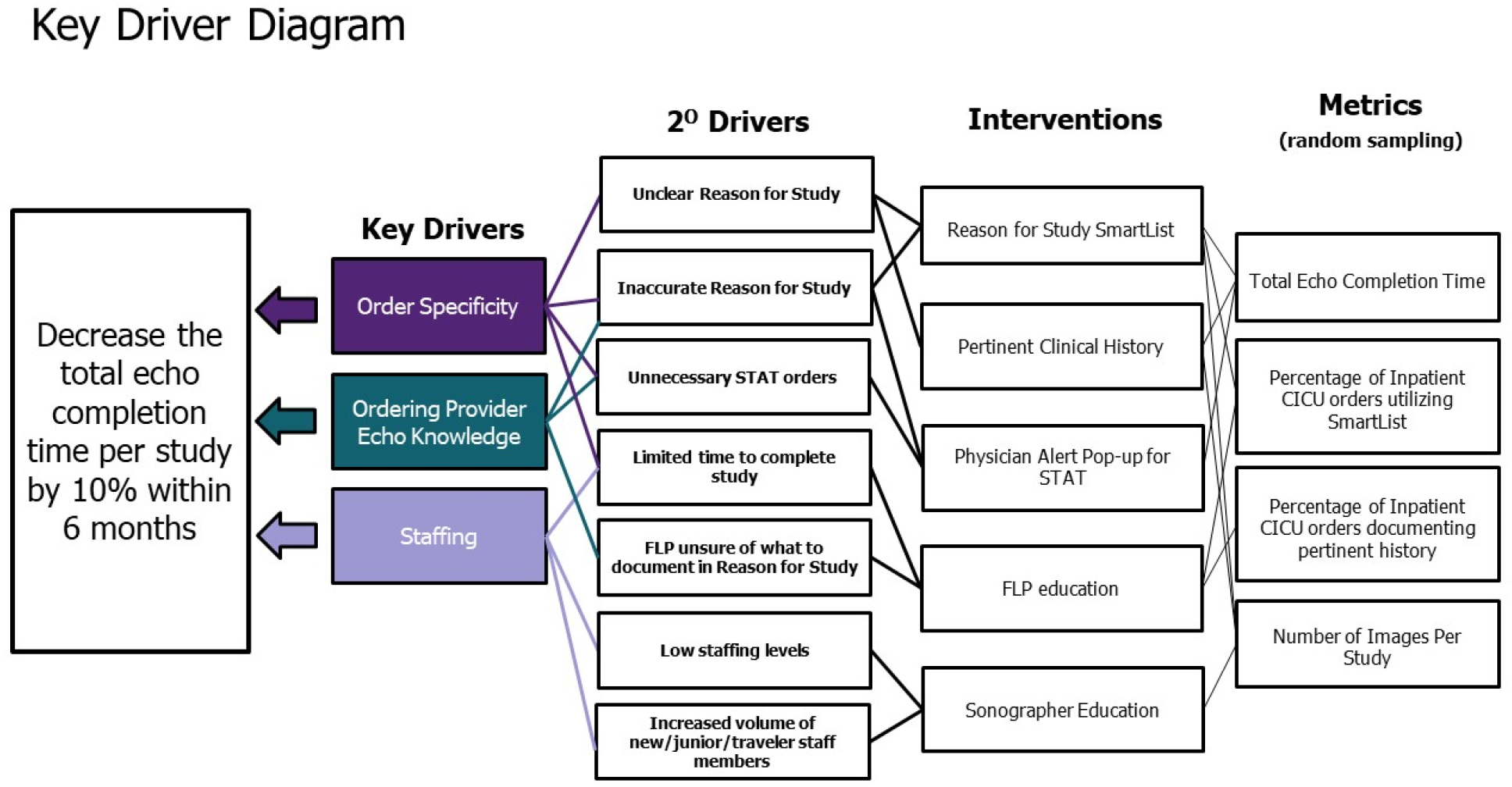
Key Driver Diagram demonstrating the SMART (specific, measurable, achievable, relevant, and time-bound) aim, key drivers, interventions, and primary measures.

**Figure 4:**
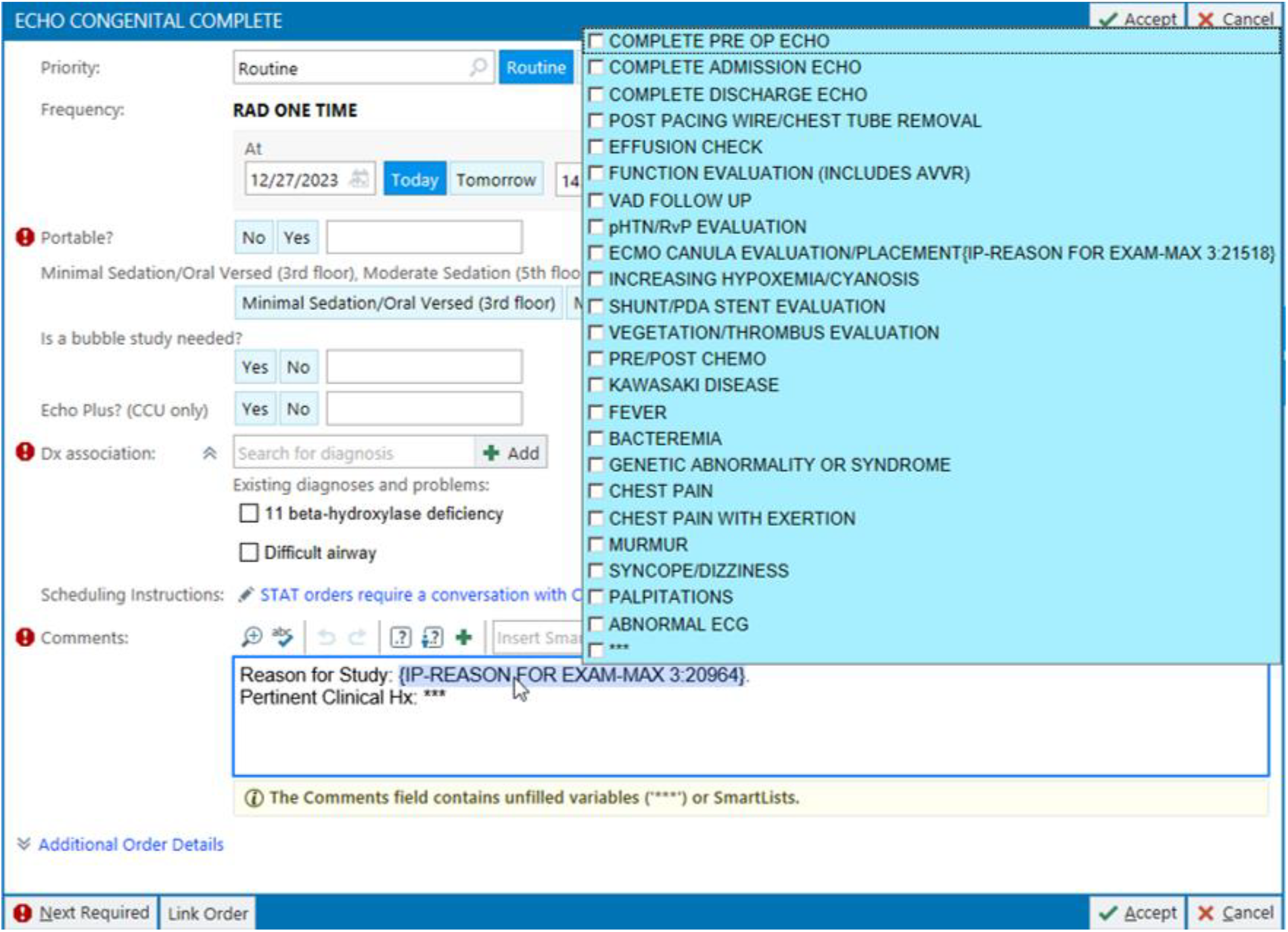
Updated inpatient echocardiogram order in Epic (Epic Systems Corporation, USA) with picklist with common reasons for ordering echocardiograms. The “ECMO” selection has an additional sub-picklist allowing for documentation of type of ECMO cannulation.

The IT team also aided in creating a provider alert which would pop-up on any STAT echocardiogram orders being signed. This showed the contact information for the inpatient echo lab as well as after-hours contacts to ensure STAT orders were appropriately communicated to facilitate rapid response. This provider alert also allowed ordering providers to go back and alter the urgency of the echo order to ASAP or Routine if the STAT order was being placed inadvertently.

Educational information was sent to the Heart Center global e-mail to update providers at all levels of these changes. The same information was also provided to the general pediatric residency group as residents were frequently the front-line providers ordering studies in clinical areas outside the CCU. Virtual meetings were held with the various Heart Center Advanced practice provider (APP) groups who work in the CCU to further review the changes and discuss the importance of specificity in the echocardiogram orders.

We performed three plan-do-study-act (PDSA) cycles to implement our interventions. Our PDSA cycles included (1) implementation of the original echocardiogram order picklist with broad e-mail notifications to the Heart Center and residency program at roll-out, (2) implementation of the STAT order physician alert, and (3) refinement of the echocardiogram order picklist, including additions for ECMO studies, and individualized educational sessions for APPs.

### Measures

Both process and outcome measures were established for the project. Measures were evaluated on complete echocardiograms performed within the CCU on weekdays in order to control for expected variation in the workflow during on-call periods at nights and weekends. Complete echocardiograms were identified by their order types: “Echo Complete Panel” and “Echo Congenital Complete”. Data collection on all measures was done retrospectively after each of the 3 PDSA cycles by a single QI team member to ensure consistency of data collection. Studies were eliminated from review if there had been patient demise or if there was extensive advanced imaging utilized such as use of Ultrasound Enhancing Agents or 3-dimensional anatomic evaluation due to their expected lengthening on the scanning time.

The primary outcome measure was the total echo completion time defined as the total amount of time necessary to complete the echocardiogram. The starting time was based upon the echocardiogram start time in Epic (Epic Systems Corporation, USA), and the ending time was determined by the timestamp on the final image in Syngo Dynamics (Siemens Healthcare, USA). The secondary outcome measure was the initial buffer time defined as the time preparing for the echocardiogram but when no active imaging had occurred. This was calculated by using the total echo completion time and subtracting the active scanning time as determined by the first and last image timestamps in Syngo Dynamics (Siemens Healthcare, USA).

Two process measures were evaluated. The number of images obtained on each echocardiogram performed in the study period. Also the percentage of echocardiogram orders documenting pertinent clinical history and/or utilizing the new order picklist in Epic (Epic Systems Corporation, USA).

Balancing measures were utilized to assess unexpected changes or other unintended consequences. This included tracking the number of inpatient echocardiograms per day during the study period to assess for any large changes in daily volumes. A follow up provider survey was also utilized to assess for CCU and echo lab staff impressions of the changes at the end of the study, including how it may have positively or negatively impacted their daily workflow.

### Analysis

We analyzed our data using run charts for both the process and outcome measures. The average of all echocardiograms reviewed at baseline and each of the 3 PDSA cycles was plotted and compared for changes over time. The percent change from baseline at the third PDSA cycle was also calculated for all measures.

Surveys were performed at baseline and after the 3^rd^ PDSA cycle. These surveys were collected via our internal Microsoft Forms SharePoint site, with links sent to all CCU attendings, CCU advanced practice providers, pediatric cardiology fellows, echo lab attendings, and echo lab sonographers.

### Ethical Considerations

The Office of Research Integrity and Compliance at Ann & Robert H. Lurie Children’s Hospital of Chicago reviewed our project prior to initiation and determined that the project did not meet the definition of human subjects research and was designed as a QI project. Therefore it did not require review by the Institutional Review Board. The physicians, advanced practice providers, and sonographers involved in this project were aware that their participation was voluntary. We used the SQUIRE (Standards for Quality Improvement Reporting Excellence) 2.0 guidelines to report our findings.^4^

## Results

We reviewed 395 echocardiograms and orders during the study period with 340 meeting criteria for final analysis. Of these studies, 88 were reviewed at baseline, 110 after PDSA cycle 1, 75 after PDSA cycle 2, and 67 after PDSA cycle 3. The average weekday inpatient echo volume remained relatively stable during the study period. The total echo completion time and calculated buffer time both decreased steadily throughout the study period for a total improvement of 25% and 56% respectively (Figure 5). During the same time period, the number of images obtained on average per echocardiogram increased. The documentation of pertinent clinical history, including the use of the order picklist, increased by 18% and maintained at 99% compliance for the final two PDSA cycles (Figure 5). For full details, see Table 1.

**Table 1:** Full results from all measures for baseline and all 3 PDSA cycles, including percentage change from baseline for the process and outcome measures.

|  | Baseline | PDSA 1 | PDSA 2 | PDSA 3 | Change from baseline |
| --- | --- | --- | --- | --- | --- |
| **Total Echo Completion Time (min)** | 68 | 61 | 54 | 51 | -25% |
| **Buffer/Waste Time (min)** | 32 | 24 | 16 | 14 | -56% |
| **Images per study** | 106 | 115 | 107 | 125 | 18% |
| **Images/min average** | 2.9 | 3.2 | 3.6 | 3.5 | 21% |
| **Documentation of pertinent clinical history (%)** | 84% | 96% | 99% | 99% | 18% |
| **Echos Reviewed** | 88 | 110 | 75 | 67 | n/a |
| **Weekday CCU volume** | 7.9 | 6.1 | 5.4 | 7.5 | n/a |
| **Weekday IP volume** | 16.5 | 18.2 | 16.7 | 17 | n/a |

**Figure 5:**
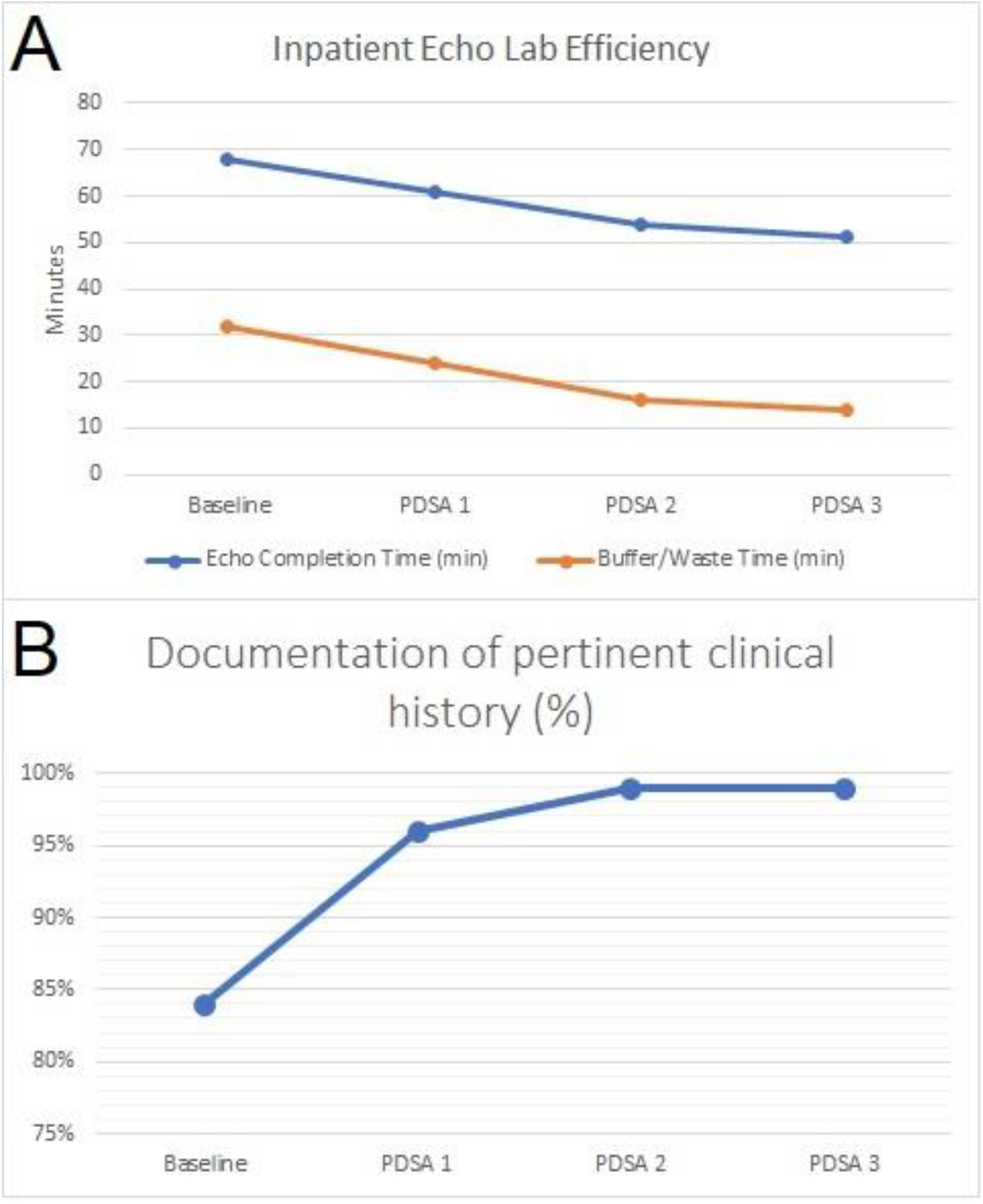
Run charts of outcome measures (A) and process measure (B) showing improvement in these measures over the course of the study period.

### Balancing Measures

A baseline provider survey was completed before the first intervention and a repeat provider survey was completed after PDSA cycle 3. 35 providers (20 echo lab providers, 15 ordering providers) completed the baseline survey and 27 providers (10 echo lab providers, 14 ordering providers) completed the final survey. Ordering providers included CCU attendings, CCU APPs, general cardiology attendings, and fellows. Echo lab providers included attending physicians and sonographers. Full details are listed in Table 2. Of note, the personnel completing the baseline and follow up surveys differed due to routine staffing changes within all departments over the course of the study period.

**Table 2:**
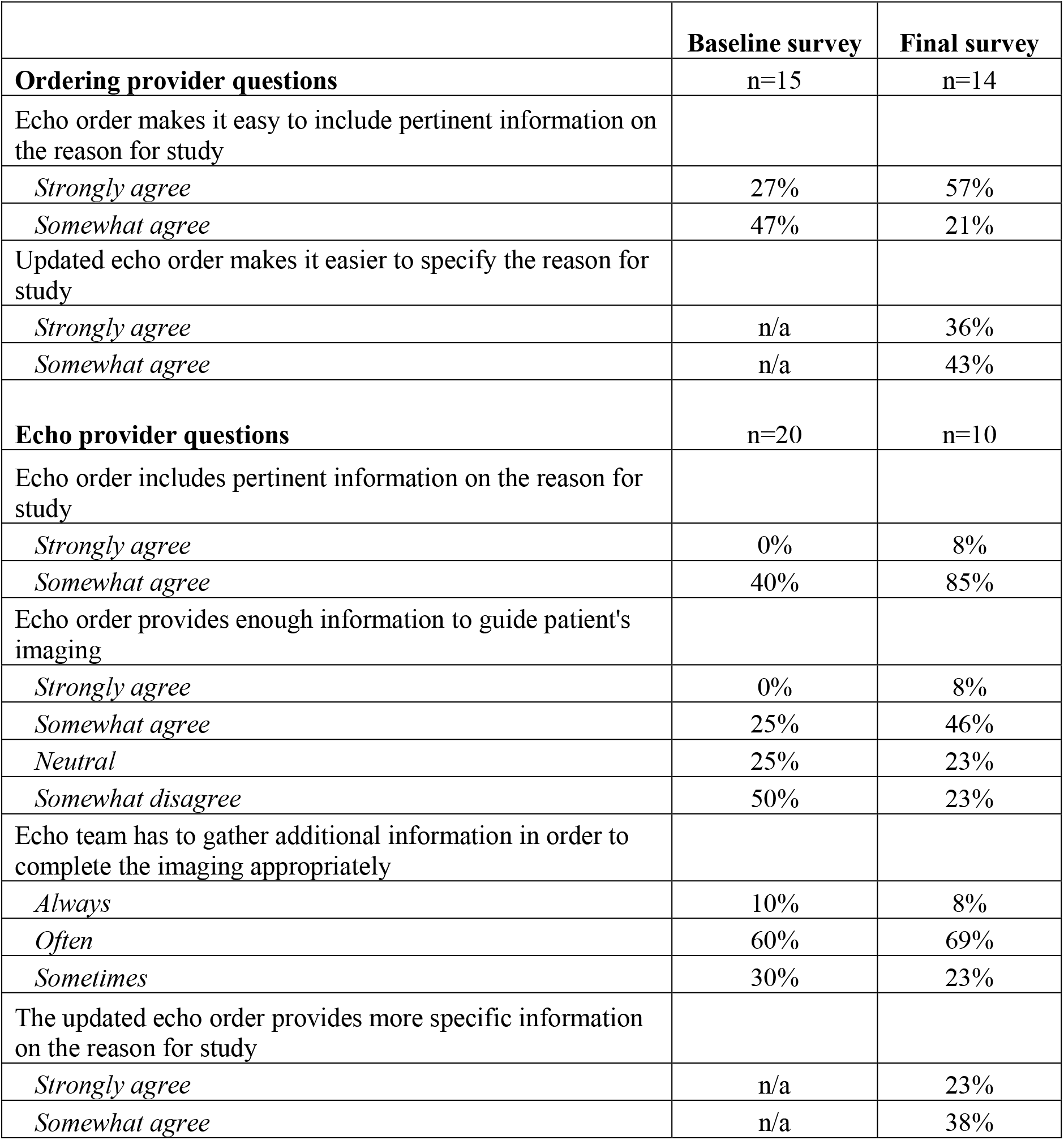
Provider survey results from baseline survey and final survey administered after PDSA cycle 3. Final survey included additional questions for balancing measures.

|  | **Baseline survey** | **Final survey** |
| --- | --- | --- |
| **Ordering provider questions** | n=15 | n=14 |
| Echo order makes it easy to include pertinent information on the reason for study |  |  |
| *Strongly agree* | 27% | 57% |
| *Somewhat agree* | 47% | 21% |
| Updated echo order makes it easier to specify the reason for study |  |  |
| *Strongly agree* | n/a | 36% |
| *Somewhat agree* | n/a | 43% |
| **Echo provider questions** | n=20 | n=10 |
| Echo order includes pertinent information on the reason for study |  |  |
| *Strongly agree* | 0% | 8% |
| *Somewhat agree* | 40% | 85% |
| Echo order provides enough information to guide patient's imaging |  |  |
| *Strongly agree* | 0% | 8% |
| *Somewhat agree* | 25% | 46% |
| *Neutral* | 25% | 23% |
| *Somewhat disagree* | 50% | 23% |
| Echo team has to gather additional information in order to complete the imaging appropriately |  |  |
| *Always* | 10% | 8% |
| *Often* | 60% | 69% |
| *Sometimes* | 30% | 23% |
| The updated echo order provides more specific information on the reason for study |  |  |
| *Strongly agree* | n/a | 23% |
| *Somewhat agree* | n/a | 38% |

Baseline survey results indicated relative satisfaction from the ordering providers on their process, but significant issues from the echo lab providers. This included 70% of echo lab providers having to always or often spend time gathering additional information before completing the imaging. When asked for feedback, one CCU attending noted “I know what I want to look at, but it is often a frontline provider placing the order who has a less robust understanding. I think check boxes will help better ensure the right things are being looked at.”

Final survey results showed an improvement in satisfaction of the ordering providers with an increase from 27% at baseline to 57% on follow up in those who strongly agreed that the echocardiogram order made it easy to include pertinent information. When asked if the order updates made this process easier, 36% strongly agreed and 43% somewhat agreed. The echo lab providers also noted improvement with the echocardiogram order including enough information to guide imaging now having 8% strongly agree (0% at baseline) and 46% somewhat agree (25% at baseline). When asked if the updated order included more specific information than the older orders had, 23% strongly agreed and 38% somewhat agreed.

## Discussion

Our study improved echo lab efficiency by implementing system changes in workflow utilizing quality improvement processes. Quality improvement processes including mechanisms for continuous quality improvement are considered standard practice in pediatric echocardiographic laboratories, including forming some of the requirements for accreditation by the Intersocietal Accreditation Commission.^5,6^ Despite this, there are limited publications addressing quality improvement initiatives within the pediatric echocardiography laboratory. Quality improvement initiatives focusing on processes rather than individuals are more likely to result in qualitative and quantitative improvements which translates to improved care for our complex patients in pediatric cardiology.^7^ In our study, we identified that inadequate echo order indications and inadequate ordering provider knowledge base were areas of improvement that we could implement to improve the efficiency of the echo lab. These changes have prompted further related QI work in our echo lab and also have the potential for reducing diagnostic errors.

Our improvement project focused on improving efficiency within our pediatric echocardiography laboratory at a busy tertiary pediatric medical center. Similar to other reports, our project demonstrates that relatively simple workflow changes can result in meaningful improvements in productivity and efficiency within the pediatric echo lab.^1-3,8^ We were able to significantly decrease the time needed for preparatory work prior to beginning the echocardiogram imaging by 56% during our study period. This decrease would provide enough time for sonographers to complete one additional echocardiogram each day, thereby reducing the number of echocardiograms delayed or requiring completion by fellows in training during their overnight call period. Similar workflow changes in other studies have demonstrated increased productivity and efficiency within the echo lab as well as decreased costs.^1^ Another study has demonstrated that echocardiogram scanning time does not correlate well with charges but does reflect resource utilization more accurately.^9^

Workflow changes can result in lasting improvements as they do not depend on ongoing education with the addition of new staff members as our project demonstrated with continued and sustained improvement over several PDSA cycles despite the addition of multiple inexperienced sonographers and new CCU staff during our study period. Considering the effectiveness and reliance on persons versus systems is important when planning QI interventions, as those dependent on systems changes are generally more successful than those solely reliant on persons.^10^ In our project, we utilized a combination of these recognized quality intervention categories in order to create meaningful change that could persist even in the face of staffing changes in the multiple departments involved in the pediatric echocardiogram process. For example, our two primary interventions would be classified as a forcing function and warning/alert respectively with the lower human reliability of education used only as a supportive measure for our other improvements. The use of interventions with higher systems reliability also helps sustaining improvements for the longer term as seen in our study.

Documenting accurate reasons for obtaining echocardiograms is also important for the pediatric echo lab as it relates to the potential for diagnostic errors. With the various and complex diagnoses pediatric echo labs routinely encounter in their daily workflow, having a misleading patient history or incorrect or no clinical question indicated on echocardiogram orders can contribute to diagnostic errors. While this accounts for only 1% of diagnostic errors, these issues can also contribute to cognitive errors for the reading physicians given the distractions it imposes, and cognitive errors can account for 37% of diagnostic errors.^11^ Thus having an accurate clinical question and patient history is critical to obtain complete and accurate imaging.

### Limitations

Our feedback from individual providers both at baseline and at the conclusion of the project was limited to those who responded to the survey. While we had responses from both CCU and echo lab staff on each survey, there could be a selection bias in those willing to complete the survey.

## Conclusion

In our improvement project, the efficiency of our inpatient echo lab significantly improved across all PDSA cycles. While these improvements occurred, there were no unintended consequences uncovered with our provider surveys, and most providers viewed the changes in a positive light. This demonstrates the ability to affect clinically relevant improvements utilizing simple workflow process changes in the pediatric echo lab. Further work is underway to improve efficiency in both the inpatient and outpatient echo labs based upon areas of concern in the original fishbone diagram. We are hopeful this work at our center can serve as a guide for other pediatric echo labs aiming to improve their daily efficiency.

## Data Availability

The data that support the findings of this study are available from the corresponding author upon reasonable request.

